# The Exposome and its Associations with Broad Mental and Physical Health Measures in Early Adolescence

**DOI:** 10.1101/2021.08.11.21261918

**Authors:** Tyler M. Moore, Elina Visoki, Stirling T. Argabright, Grace E. DiDomenico, Ingrid Sotelo, Jeremy D. Wortzel, Areebah Naeem, Ruben C. Gur, Raquel E. Gur, Varun Warrier, Sinan Guloksuz, Ran Barzilay

**Author notes:** Correspondence concerning this article should be addressed to Ran Barzilay, Perelman School of Medicine, University of Pennsylvania, 3400 Spruce St. – 10th Floor Gates Pavilion, Philadelphia, PA 19104.

## Abstract

Exposures to perinatal, familial, social, and physical environmental stimuli can have substantial effects on human development. Yet the complex network structure of the environment (i.e., exposome) makes it challenging to investigate. Here, we analyze the exposome using data from the Adolescent Brain Cognitive Development Study (ABCD, N = 11,235, mean age = 10.9, 52% male) and replicate key findings in an age and sex matched sample from the Philadelphia Neurodevelopmental Cohort (PNC, N = 4,993). Both these cohorts are large, diverse samples of US adolescents with phenotyping at multiple levels of environmental exposure. In ABCD, applying data-driven iterative factor analyses and bifactor modeling, we reduced dimensionality from n=798 exposures to six exposome subfactors and a general (adverse) exposome factor. These factors revealed quantitative differences among racial and ethnic groups. Exposome factors increased variance explained in mental health by 10-fold (from <4% to >38%), over and above other commonly used sociodemographic factors. The general exposome factor was associated with psychopathology (β=0.28, 95%CI 0.26-0.3) and key health-related outcomes: obesity (OR=1.4, 95%CI 1.3-1.5) and advanced pubertal development (OR=1.3, 95%CI 1.2-1.5). In PNC, using substantially fewer available environmental exposures (n=29), analyses yielded consistent associations of the general exposome factor with psychopathology (β =0.15, 95%CI 0.13-0.17), obesity (OR=1.4, 95%CI 1.3-1.6) and advanced pubertal development (OR=1.3, 95%CI 1-1.6). Findings demonstrate how incorporating the exposome framework can be useful to study the role of environment in human development.

---

Environment (E) is a key driver of variability in human development^1^, with extensive literature linking environment to general^2^ and mental health^3^. Childhood environment is especially important for development, with evidence that exposures occurring during sensitive periods of development are critical for later life health outcomes in both animals^3^ and humans^4^. Therefore, there is a clear need to characterize environment in a systematic and comprehensive manner early in the lifespan to advance our understanding of its role in human development.

Three major challenges in studying environment’s associations with health and disease are notable. First, exposures are often co-occurring and collinear^5^, and it is difficult to disentangle specific effects because they are intertwined in a complex, dynamic network^6^. For example, when studying exposure to trauma, one should consider its correlation with, for example, poverty, neighborhood environment, and familial factors^7^. Thus, it is difficult to dissect specificity in relationships between single exposure types (e.g., trauma) and developmental outcomes. Second, exposures are not isolated and are likely to interact both among themselves (ExE) and with genetics (G) (GxE) to drive developmental outcomes, as proposed in various developmental models (e.g., “stress-diathesis”^8^, “stress inoculation”^9^, “developmental origins of health and disease”^10^). Finally, for complex conditions it is exceptionally difficult to clearly separate exposures into genetic and environmental influences as the environment is reflected in genetic association studies and genetics shape our environment^11–15^. Hence, categorizing variables as purely biological or environmental is impossible.

Specifically for the first challenge of collinearity, the exposome paradigm is one framework that may advance the study of environment^16^. The exposome (see Wild 2005^17^) represents the totality of environmental exposures that an individual experiences from conception throughout the lifespan^18, 19^, as well as the interaction among these exposures^6^.

Though early studies of the association between exposome and health were focused on physical exposures (e.g., chemical carcinogens) on cancer risk^20^, the concept has been extended to include environmental exposures at a broader context including socioeconomic and lifestyle factors^21^.

More recently, the exposome framework has been applied in psychiatry^22^, with evidence of exposome effects in both psychosis^23, 24^ and suicide research^25^.

While associations of specific environmental exposures and development have long been studied^26^, there is a need for an integrative approach that can leverage environmental exposures’ data to generate measures that will capture the main components of the exposome comprehensively, test its relationship with health measures, and facilitate integration of exposome measures in studies of human development. Specifically, there is a gap in large-scale studies on the association between the exposome and child and adolescent development. The availability of rich data on many levels of environmental exposures in youth cohorts provides an opportunity to address this gap. Here, we apply an exposome framework analysis in two youth datasets - The Adolescent Brain and Cognitive Development (ABCD) Study^27^ and the Philadelphia Neurodevelopmental Cohort (PNC)^28^.

The ABCD follows a large, diverse cohort of children (N=11,878, recruited at age 9-10) ascertained through school systems and spanning almost the entire geographic United States, including both urban and rural settings^27^. ABCD Study protocol collected data on environment at multiple levels of exposure including household, family, school, neighborhood, and state-level^29^. Several hypothesis-driven studies have examined specific ABCD exposures’ effects on brain and behavior outcomes (e.g., trauma^30^, neighborhood poverty^31^, air pollution^32^, prenatal cannabis exposure^33^, screen time^34^, family factors^35^). In the current work, we employed an exposome framework approach that systematically investigates multiple environmental exposures. We used data from the 1-year follow-up ABCD Study assessment (N=11,235, see **Supplemental Table 1** for demographics), which included youth- and parent-report of children’s exposures and census- level data^29^. We conducted a series of factor analyses to reduce the dimensionality of the data and generate exposome factor scores. Additionally, we aimed to test the external validity of our exposome conceptual framework using an independent age and gender matched sample (N=4,993) of American youth from the PNC ^28^. While PNC was less focused on environment compared to ABCD and did not collect data on school and family dynamics, it still included a few measures on environmental exposures (e.g., trauma^36, 37^, neighborhood level socioeconomic factors^38^) based on youth and parent report and geocoded census-level data, allowing us to apply an exposome approach analyses in PNC in an attempt to generalize findings obtained in ABCD.

In view of the exposome paradigm that multiple environmental exposures are associated with the variability in health outcomes, we aimed to (i) comprehensively and systematically characterize the exposome (i.e., the combined effect of exposures at multiple levels of analysis) of early-adolescents in the US using two youth cohorts; (ii) generate exposome scores that represent environment and can be used for downstream analyses; and (iii) test exposome’s associations with mental health and indicators of general health, over and above commonly used proxies of socioeconomic environment (parent education and household income). For general health outcomes, we focused on obesity, a key risk factor for later lifespan morbidity^39^, and pubertal development, considering studies linking earlier puberty with poorer health outcomes^40^. **Figure 1** depicts the overall study design.

**Figure 1.**
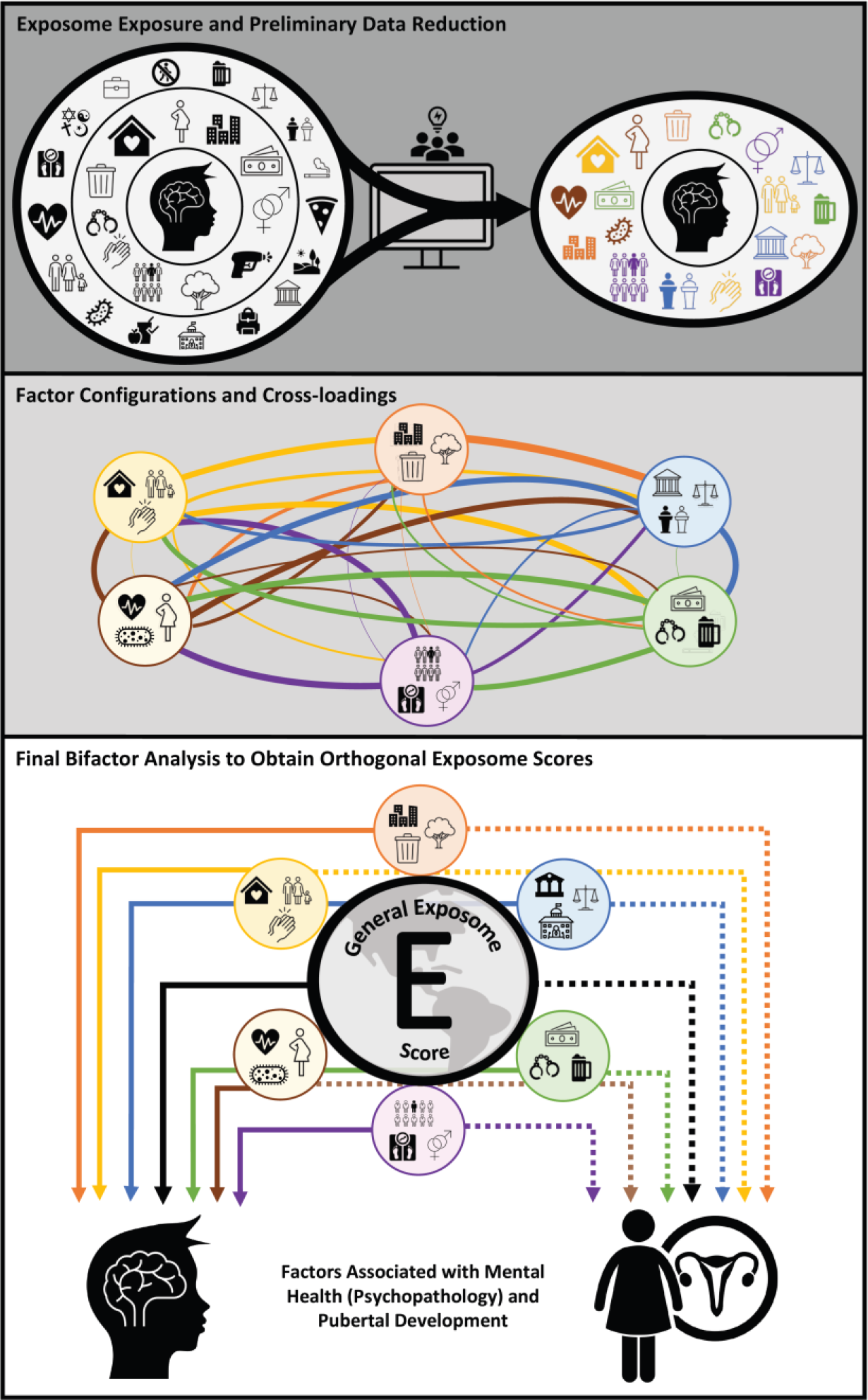
Visual presentation of study design. First, 798 environmental variables from the ABCD Study were chosen for representing the multiple dimensions of the exposome. These variables were reduced to 348 variables based on choices to use ABCD Study’s summary measures, and then further reduced using an iterative process of exploratory factor analyses that identified correlated factors allowing reduction to 96 variables from multiple dimensions of environment including family, household, school, extracurricular, neighborhood and state-level and prenatal and history of antenatal exposures. (top panel). Thereafter, these 96 combined items underwent an exploratory factor analysis that culminated in a final model, which finalized factor configurations and cross-loadings (middle panel), revealing 6 factors relating to the exposome (household adversity factor, neighborhood environment factor, day-to-day experiences factor, state conservatism-ruralness factor, family values factor, and pregnancy/birth complications factor). Subsequently, these factors were subjected to confirmatory bifactor analysis, which allowed the generation of a general exposome factor informed by all items, in addition to six orthogonal exposome subfactors (bottom panel). Finally, we investigated how these exposome factors are associated with mental health, body mass index, and pubertal (pre-)development.

## Results

### Dimensionality reduction of the exposome in the ABCD Study

We identified a comprehensive set of environmental exposures in the ABCD Study (798 variables, **Supplemental Table 2**). In line with our goal to comprehensively assess environment, and the conceptual exposome framework that multiple exposures combine to explain variance in health outcomes, and because genetics and environment inform and interact with each other, we applied a permissive definition of environment using all available data on environment as collected by multiple sources (i.e., youth-reported, parent-reported, and census derived environmental variables). For example, since parental factors play a major role in childhood development, we included parental psychopathology in our analyses, even though we acknowledge that genetic contributions of parental psychopathology also exist in the child.

Furthermore, because we wanted to investigate the utility of applying an exposome framework, we excluded two pivotal measures commonly used to estimate environment, including in previous ABCD Study research: household income^41, 42^ and parental education^43^. This choice allowed us (1) to test the “added value” of the exposome scores to explain variance in health outcomes over and above commonly used proxies of environment known to associate with developmental outcomes^44^, including in ABCD Study^42^; and (2) to validate the exposome scores using “classic” indicators of socioeconomic environment.

From the 798 identified environmental variables, we decided on features for which to use ABCD summary measures (e.g., family conflict; see detailed description of variable choice in **Methods**), resulting in 348 variables for analysis. Then, we applied a set of exploratory factor analyses (EFAs) to identify correlation-based clustering among variables and allow further reduction of variable number. **Supplemental Figure 1** provides a schematic presentation of this dimensionality reduction process, which is described in full in **Methods**. Briefly, we started by including all 348 variables in analysis and, using nine EFAs, iteratively reduced these to 96 with minimal redundancy. Each of the EFAs described above included items from subdomains of environmental exposures, including parental mental health and drug use (**Supplemental Table 3**), maternal substance use during pregnancy (**Supplemental Table 4**), neighborhood-level characteristics (**Supplemental Table 5**), household-level poverty and religiosity (**Supplemental Table 6**), school-level characteristics (**Supplemental Table 7**), pregnancy complications (**Supplemental Table 8**), birth complications (**Supplemental Table 9**), parent-report of childhood traumatic events (**Supplemental Table 10**), and youth-report of life events (**Supplemental Table 11**).

**Table 1** shows the results of the final EFA of the minimally redundant 96 environmental variables, using iterated target rotation (ITR) designed to detect complex structure (cross-loadings), which revealed six factors. *Factor 1* comprises variables most related to *household adversity*, based primarily on parent-report, with the strongest indicators being the mother’s use of tobacco or marijuana during pregnancy, parental alcohol-related problems affecting ability to hold a job or stay out of jail, and frequent adult conflict in the house. *Factor 2* comprises variables most related to *neighborhood environment*, based primarily on geocoded address, with the strongest indicators being census-derived measures of neighborhood poverty and population density. *Factor 3* comprises variables most related to youth-reported *day-to-day experiences*, both positive (e.g., feeling “involved at” and enjoying school, acceptance by caregivers) and negative (e.g., experiences of discrimination, family conflict). *Factor 4* comprises variables most related to *state environment* (i.e., environmental factors from the state-level), with the strongest indicators being negative attitudes toward persons with non-hetero sexual orientation, traditional views about the roles of women, and less permissive marijuana laws. Note that a “ruralness” aspect of *Factor 4* is evident in the low neighborhood wealth and property values (seventh indicator from top). *Factor 5* comprises variables most related to *family values*, with the strongest indicators being the strictness of rules related to alcohol, tobacco, and marijuana, as well as various indicators that tap importance of religion and family cohesiveness. *Factor 6* includes variables most related to *pregnancy and birth complications*, with the strongest indicator being premature birth. Of note, prenatal exposure to substances did not load on *Factor 6*, but rather on *Factor 1* which taps household adversity. This configuration was used because it indicates that maternal substance use is more revealing of household adversity than of pregnancy or birth complications. Inclusion of maternal substance use in *Factor 6* would, paradoxically, increase the ambiguity of that factor.

**Table 1.**
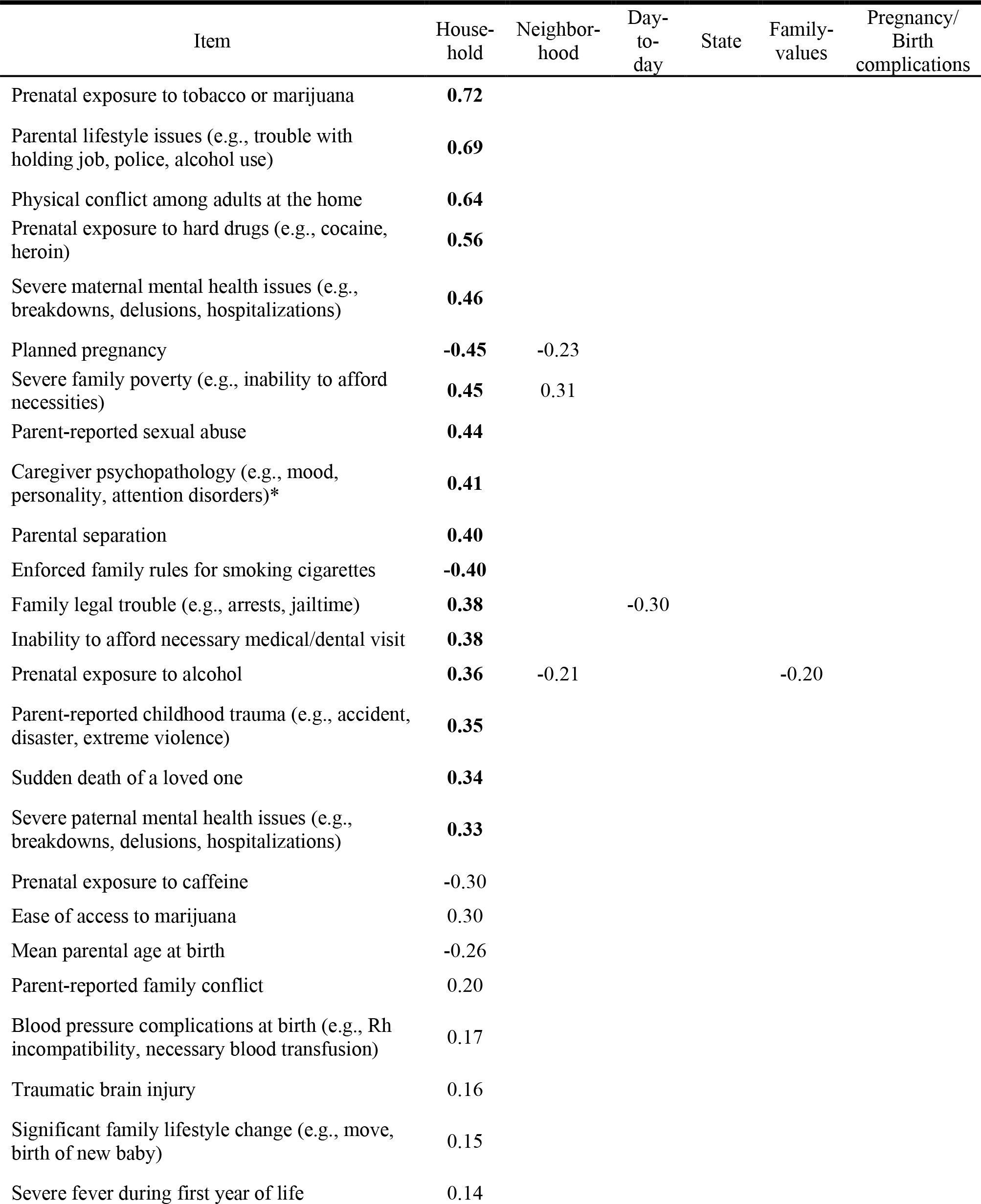
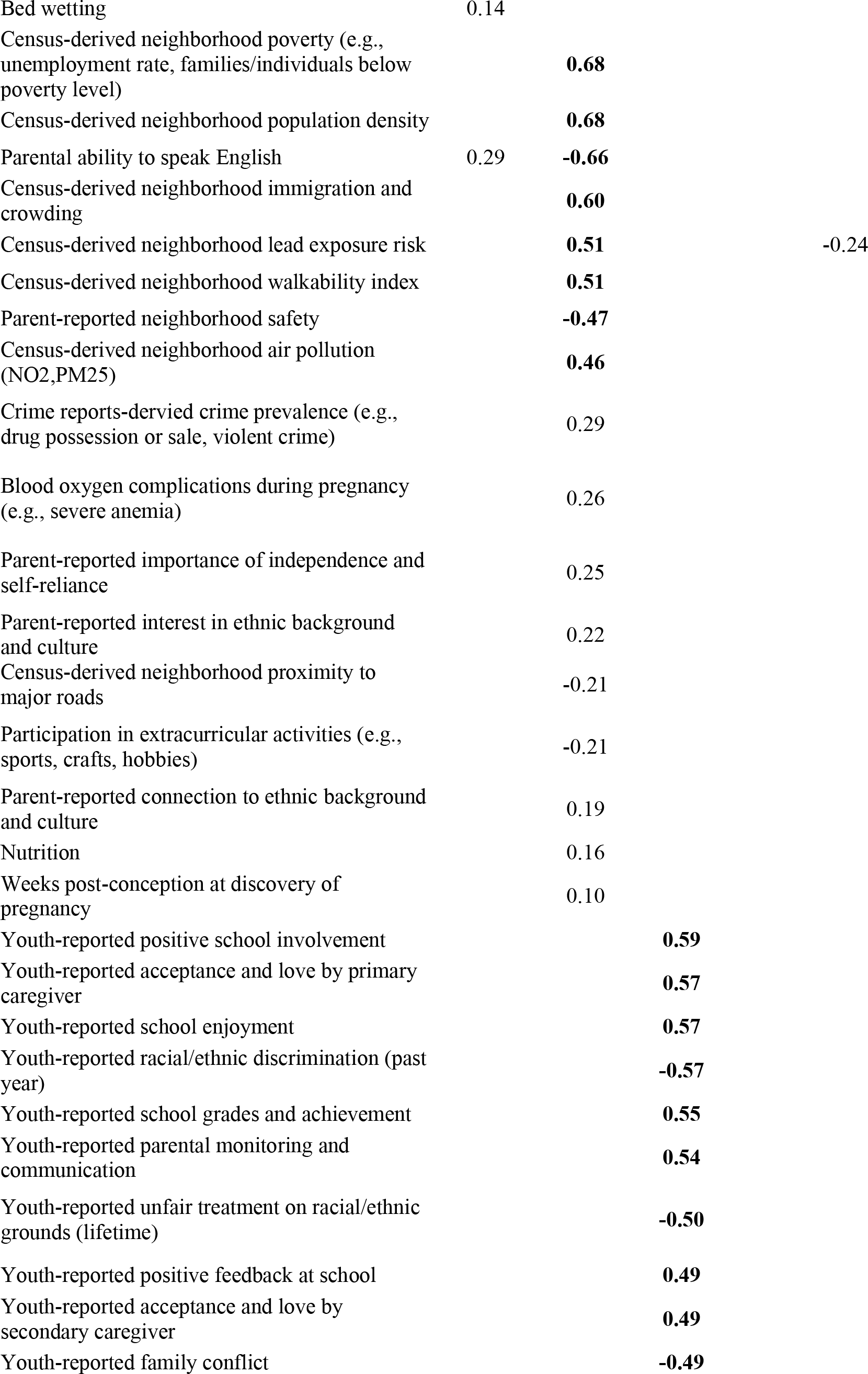
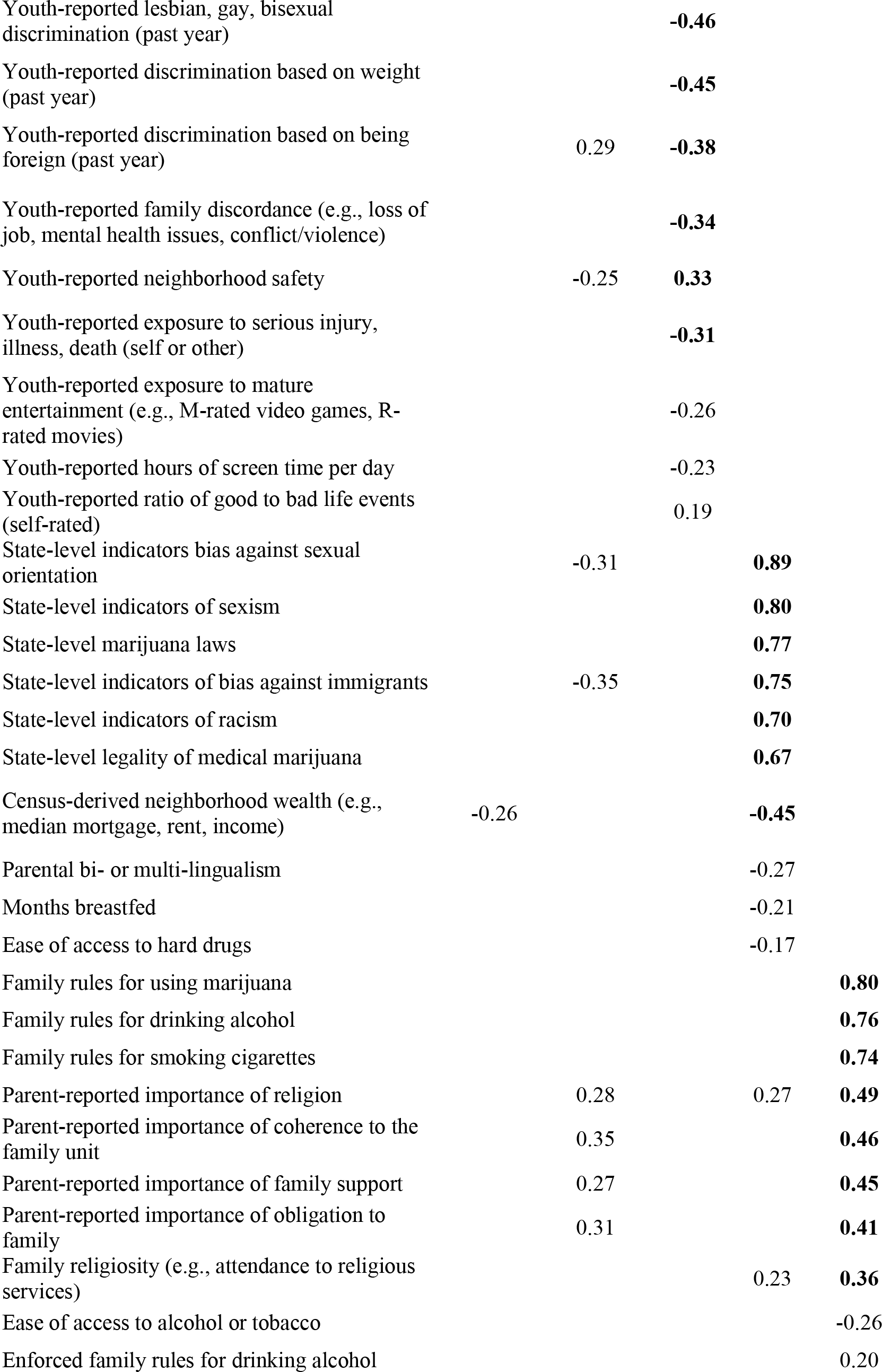
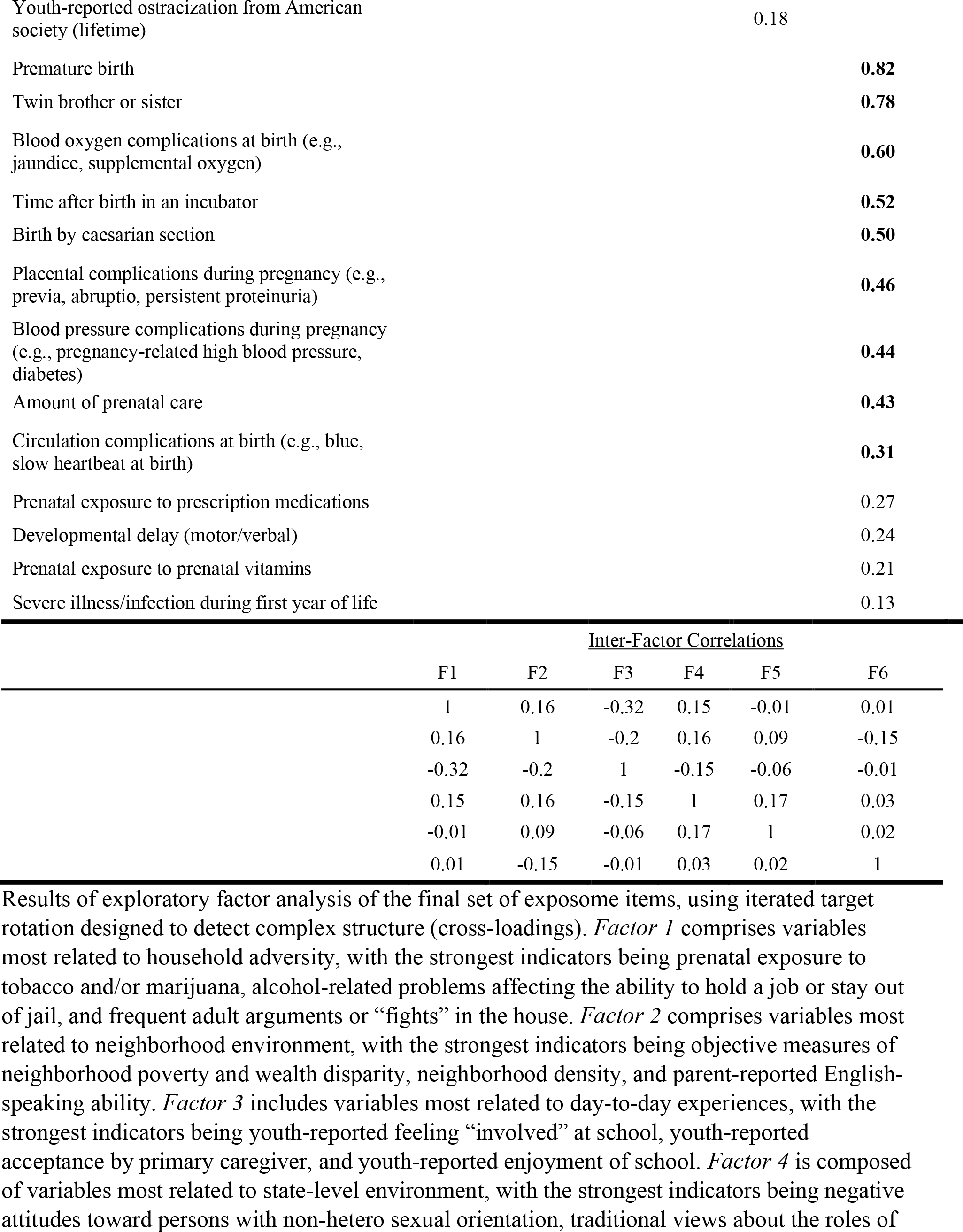
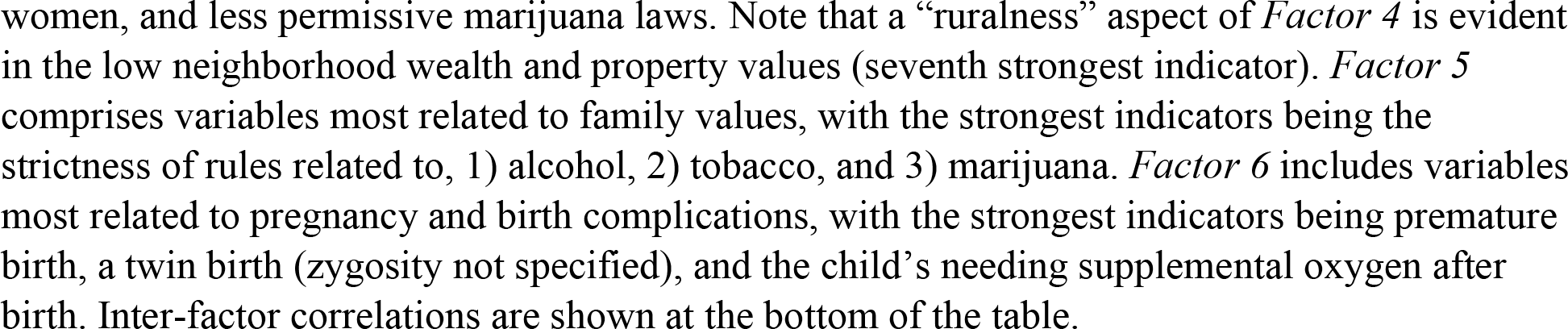
Exploratory factor analysis of the optimized collection of exposome items in the ABCD Study using iterated target rotation

We then conducted a quasi-confirmatory factor analysis (CFA) model that included 65 items, selected from the 96 variables based on their loadings in the EFA (“quasi-” because there is no cross-validation being performed here; the “confirmatory” model is actually being used to estimate a model for score creation rather than truly confirm a theoretical or empirically-derived model)^45^. A notable reason a quasi-confirmatory approach was used here is that exploratory bifactor rotations still need further development^46^. The commonly used Schmid-Leiman performs well, but it is not a true bifactor^47^. Others have not been well-tested for the use of score-creation^48, 49^. The quasi-CFA approach allows one to stay as close as possible to a true bifactor (albeit here with cross-loadings) to take advantage of its lack of proportionality constraints (as in the Schmid). This lower parsimony of a true bifactor allows one to estimate especially precise sub-factor scores^50^.

Items with absolute value less than 0.30 in the ITR-rotated EFA (**Table 1**) were removed for the final CFA analysis used for creation of exposome scores. These selected 65 items inform the resultant general exposome factor and were derived from multiple scales of the ABCD Study, from both parent- and youth-report and from census-derived measures.

### Generation of exposome scores in ABCD Study

To estimate a general exposome factor *(Exp-factor)* score and orthogonal exposome subfactor scores that allow delineation of discrete environmental effects on development, we applied a bifactor modeling approach^51^. **Figure 2** shows the results of the quasi-confirmatory bifactor analysis with the loadings of the strongest items and their direction (see full list of item loadings in **Supplemental Table 12)**. Fit of the model was acceptable^52, 53^, with a root mean-square error of approximation (RSMEA) of 0.033 and standardized root mean-square residual (SRMR) of 0.060; confidence intervals around the RMSEA were imperceptibly narrow at this sample size. Note that the comparative fit index (CFI) of 0.85 was below the acceptable range, conflicting with other fit indices, which is a known phenomenon in large models^54^ and likely does not indicate poor fit^55^. Here, it was possible to achieve a CFI > 0.90 *post hoc* by allowing some residuals to correlate, but we opted to leave the model “pure” rather than use modification indices^56^ merely to increase one fit index. Thus, the *Exp-factor* captures the broad, multidimensional environmental phenotyping of the ABCD assessment. Notably, extreme household poverty, parental legal trouble, unplanned pregnancy, physical conflict among adults in the household, neighborhood poverty, and experiences of discrimination were among the strongest loading items of the *Exp-factor*. Also, of note, in the EFA model (**Table 1**), experiences of discrimination loaded strongly on the *day-to-day experiences factor*, but in the bifactor model (**Figure 2**; **Supplemental Table 12**), variance explained in the discrimination items “shifted” from *day-to-day experiences* to the *Exp-factor*. Thus, in the final model, most discrimination is accounted for by the *Exp-factor* score. The *day-to-day experiences* subfactor is left without discrimination and is heavily influenced by attitudes toward school, a center-point of life in this age range.

**Figure 2.**
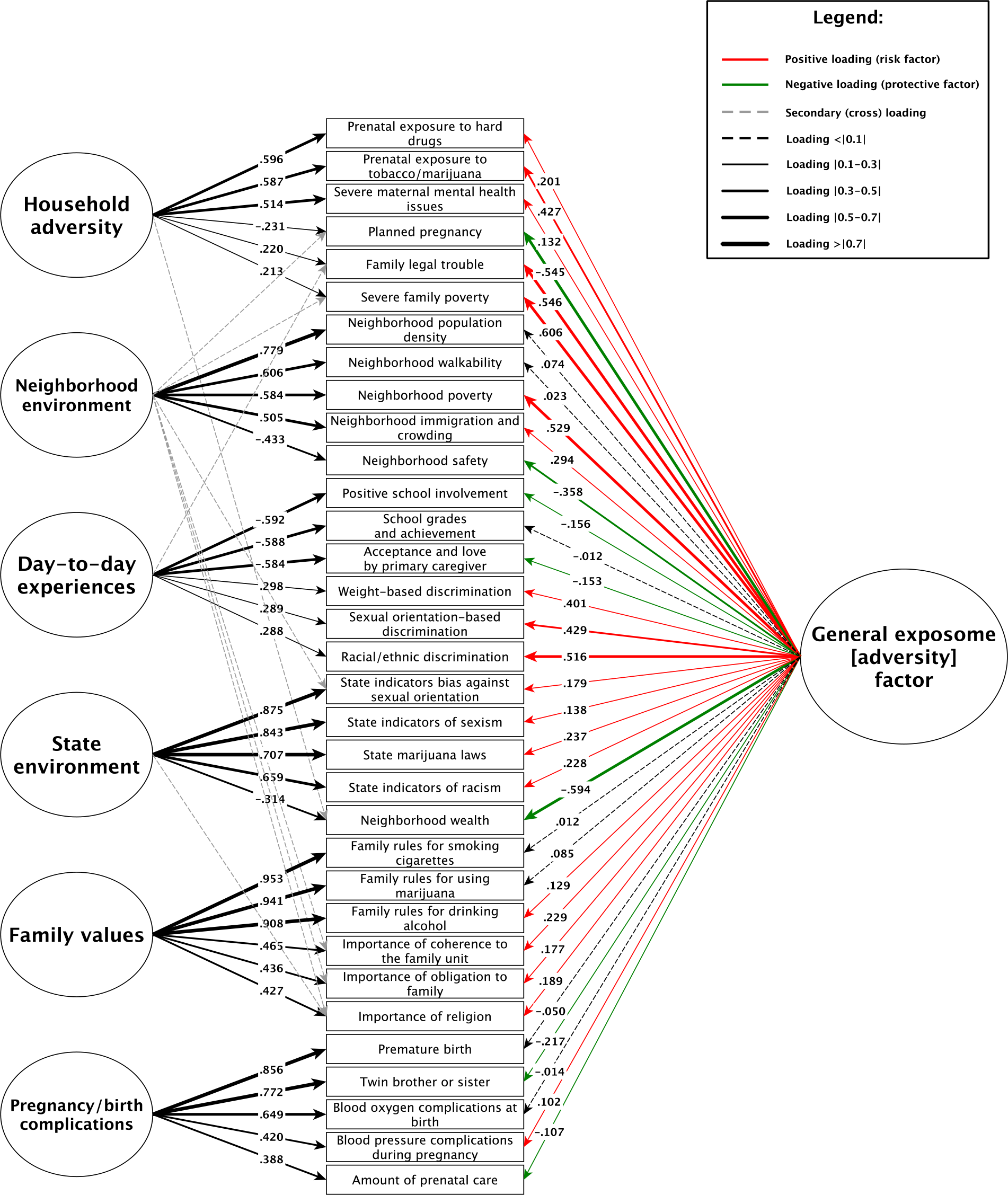
Exposome bifactor model of the ABCD Study Bifactor model of confirmatory factor analysis. Only the top 3 items loading within-factor and on the *Exp-factor* are included; that is, a specific factor’s indicators were included in the diagram if they were among the top three strongest-loading items on that specific factor *or* on the general factor (so maximum possible = 6 indicators per factor in the diagram). Arrow thickness relates to the strength of the loading (higher the loading, thicker the arrow). Arrow color relates to the sign of the loading – a red arrow corresponds to positive loading (associated with a higher *Exp-factor* score; risk factor) and a green arrow corresponds to negative loading (associated with a lower *Exp-factor* score; protective factor). Subfactors are presented from top to bottom in order from F1 to F6. See **Supplemental Table 12** for the full list of items and their loadings, and for the breakdown of variables that make up each factor in the bifactor model.

### The exposome across sociodemographic groups in ABCD Study

Next, we tested the associations of the *Exp-factor* and the six exposome subfactor scores with key sample demographics. **Figure 3** shows comparisons of the exposome scores across sex, household income, parental education, race, and ethnicity. Sex differences did not emerge in the *Exp-factor* or in five of the six subfactors; the only difference was that males had greater *day-to- day experiences* scores (Cohen’ *d*=0.30, P<.001), which is driven by the fact that males report disliking school more often than females do. Comparison of high to low parent education and household income revealed expected differences, whereby both were associated with greater *Exp-factor* score with very large effect sizes (for income, *d*=1.40; for parent education, *d*=1.16, P’s<0.001), and greater *neighborhood environment* (poverty) scores with medium effect size (for income, *d*=0.63; for parent education, *d*=0.41, P’s<0.001). Comparison of high/low parent education and income of other exposome factors including *household adversity*, *family values*, and *state environment* revealed differences in the small effect size range (*d*’s ranging from 0.10-0.22, all P’s<0.001). Notably, comparing high/low income and parent education revealed either very small (d’s<0.09), or non-significant differences in the *day-to-day experiences* subfactor and the *pregnancy/birth complications* subfactor (**Figure 3**).

**Figure 3.**
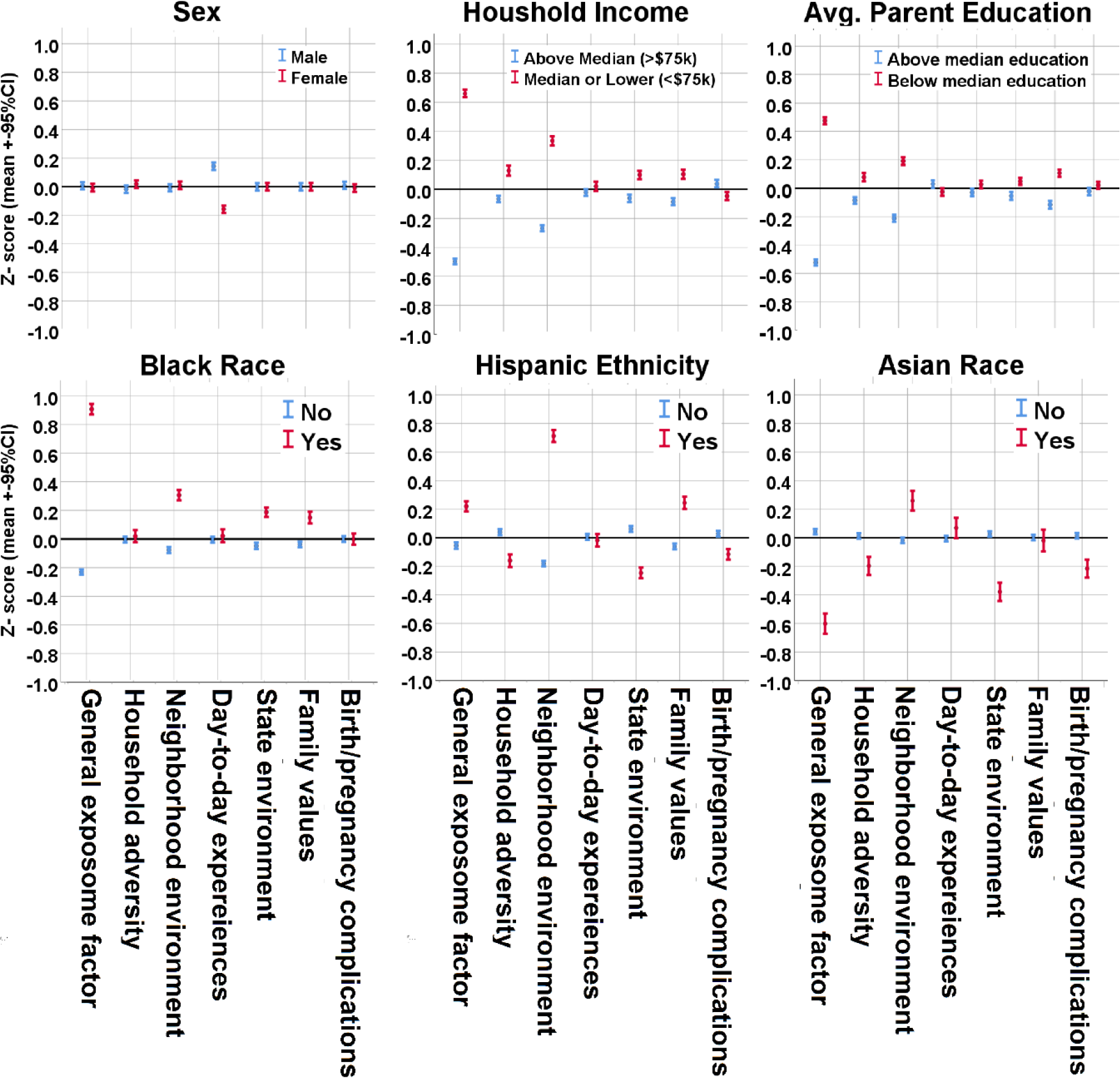
Exposome scores across demographic comparisons in the ABCD Study Exposome scores for the six orthogonal subfactors and one general factor are compared across demographic groups. Displayed are differences between male and female participants, high and low household income, and high and low parent education **(top panel)**, Black race, Hispanic ethnicity, and Asian race **(bottom panel)**. Demographic differences serve as an initial validation for use of generated exposome factor scores.

Comparison of the *Exp-factor* score across races and ethnicities revealed substantial differences. Black participants (n=2,269) had greater *Exp-factor* scores than non-Black participants (n=8,966) in the very large effect size range (*d*=1.28, P<0.001); Hispanic participants (n=2,226) also showed greater *Exp-factor* scores than non-Hispanic participants (n=8,872), but with a smaller effect size (*d*=0.29, P<0.001). Notably, Asian participants (n=723) had lower *Exp-factor* scores than non-Asian participants (n=10,512), with a medium to large effect size (*d*=0.66, P<0.001). Comparisons of exposome subfactors across races and ethnicities showed that the only difference with a large effect size was observed in Hispanic participants, who had a greater *neighborhood environment* subfactor score (representing greater population density and, to a lesser extent, poverty) (*d*=0.92, P<0.001). Similarly, Black and Asian participants showed greater *neighborhood environment* subfactor scores, but with smaller effect sizes (for Black, *d*=0.41; for Asian, *d*=0.28, P’s<0.001). Comparison of the *state environment* subfactor revealed differences among races and ethnicity at the small to moderate effect size range (*d*’s ranging from 0.25-0.43). Differences in *family values* subfactor scores were observed among Black and Hispanic, but not Asian participants, who were the only group that showed differences in the *birth/pregnancy complications* subfactor, with lower scores. Notably, no differences were observed in *day-to-day experiences* (largely determined by attitudes toward school) when comparing across races and ethnicities. (**Figure 3**).

### Association of exposome scores with mental health in ABCD Study

We next sought to use exposome factor scores to explain variance in participant mental health. First, we calculated a single general factor score that represents the overall liability to psychopathology (*P-factor*)^57, 58^, which was consistently shown to accurately represent psychopathology in youth samples^59^. Then, we used the exposome scores as independent variables to test their contribution to explaining variance in *P-factor* (dependent variable). We found that while age, sex, race, ethnicity, household income and parent education explained <4% of the variance in *P-factor* score, the addition of the exposome factors increased the variance explained ∼10 fold to 38.2% (**Table 2**). Among the exposome factors, *day-to-day experiences* showed the greatest association with *P-factor* score (Standardized Beta=0.516, P<0.001), followed by the *Exp-factor* (Standardized Beta=0.276, P<0.001). Other exposome subfactors were also significantly associated with *P-factor* score, but with relatively modest effect sizes (all betas<0.09, all P’s<0.025). The single subfactor not associated with *P-factor* score was *pregnancy/birth complications* (P=0.075).

**Table 2.** Association of exposome factor scores to psychopathology *P-factor* score in the ABCD Study

|  | Model 1 (Demographics) |  |  | Model 2 (Demographics + Exposome) |  |  |
| --- | --- | --- | --- | --- | --- | --- |
|  | Beta | SE | P | Beta | SE | P |
| Age (months) | -0.009 | 0.010 | 0.363 | 0.007 | 0.008 | 0.383 |
| Female sex | -0.098 | 0.009 | <0.001 | -0.017 | 0.008 | 0.031 |
| White race | 0.020 | 0.014 | 0.143 | -0.012 | 0.011 | 0.277 |
| Black race | 0.075 | 0.013 | <0.001 | -0.014 | 0.011 | 0.224 |
| Asian race | -0.012 | 0.008 | 0.232 | 0.001 | 0.007 | 0.870 |
| Hispanic ethnicity | 0.007 | 0.010 | 0.491 | 0.033 | 0.009 | <0.001 |
| Parent education (years) | -0.057 | 0.013 | <0.001 | -0.002 | 0.011 | 0.852 |
| Household income (ordinal) | -0.097 | 0.013 | <0.001 | 0.035 | 0.012 | 0.003 |
| General exposome adversity |  |  |  | 0.285 | 0.011 | <0.001 |
| Household adversity |  |  |  | 0.083 | 0.008 | <0.001 |
| Neighborhood environment |  |  |  | -0.021 | 0.009 | 0.024 |
| Day-to-day experiences |  |  |  | 0.518 | 0.008 | <0.001 |
| State environment |  |  |  | 0.027 | 0.008 | 0.001 |
| Family values |  |  |  | -0.019 | 0.008 | 0.018 |
| Pregnancy/birth complications |  |  |  | 0.014 | 0.008 | 0.075 |
| Adjusted R<sup>2</sup> | 0.039 |  |  | 0.382 |  |  |

Effect sizes (standardized betas) derived from a linear regression model testing association of demographics and exposome factors with general psychopathology (P-factor). Abbreviations: SE = standard error.

### Association of exposome scores with youth obesity and pubertal development in ABCD Study

Lastly, we tested whether exposome scores are associated with general adolescent-health indicators that are important for health later in the lifespan: obesity^39^ and pubertal development^40^, which are both influenced by the environment^60, 61^. Overall, 1,871 (16.7%) in the cohort were obese based on U.S. Centers for Disease Control (CDC) definitions (body mass index [BMI]>95^th^ percentile)^62^. 727 youths (6.5% of sample, n=104 males [1.7% of males], n=623 females [11.5% of females]) were late/post-pubertal (4/5 on a 5-point Likert scale). The *Exp- factor* was significantly associated with obesity and with late/post-pubertal stage (odds ratio [OR]=1.41, 95%CI=1.31-1.52; OR=1.30 95%CI=1.16-1.47, respectively, P’s<0.001; **Figure 4** and **Supplemental Tables 13-14,** models co-varied for demographics, household income, and parental education, and BMI in the puberty model). No exposome subfactors were associated with obesity. The *day-to-day experiences* subfactor was the only one significantly associated with late/post-pubertal developmental stage (OR=1.31, 95%CI=1.19-1.43, P<0.001).

**Figure 4.**
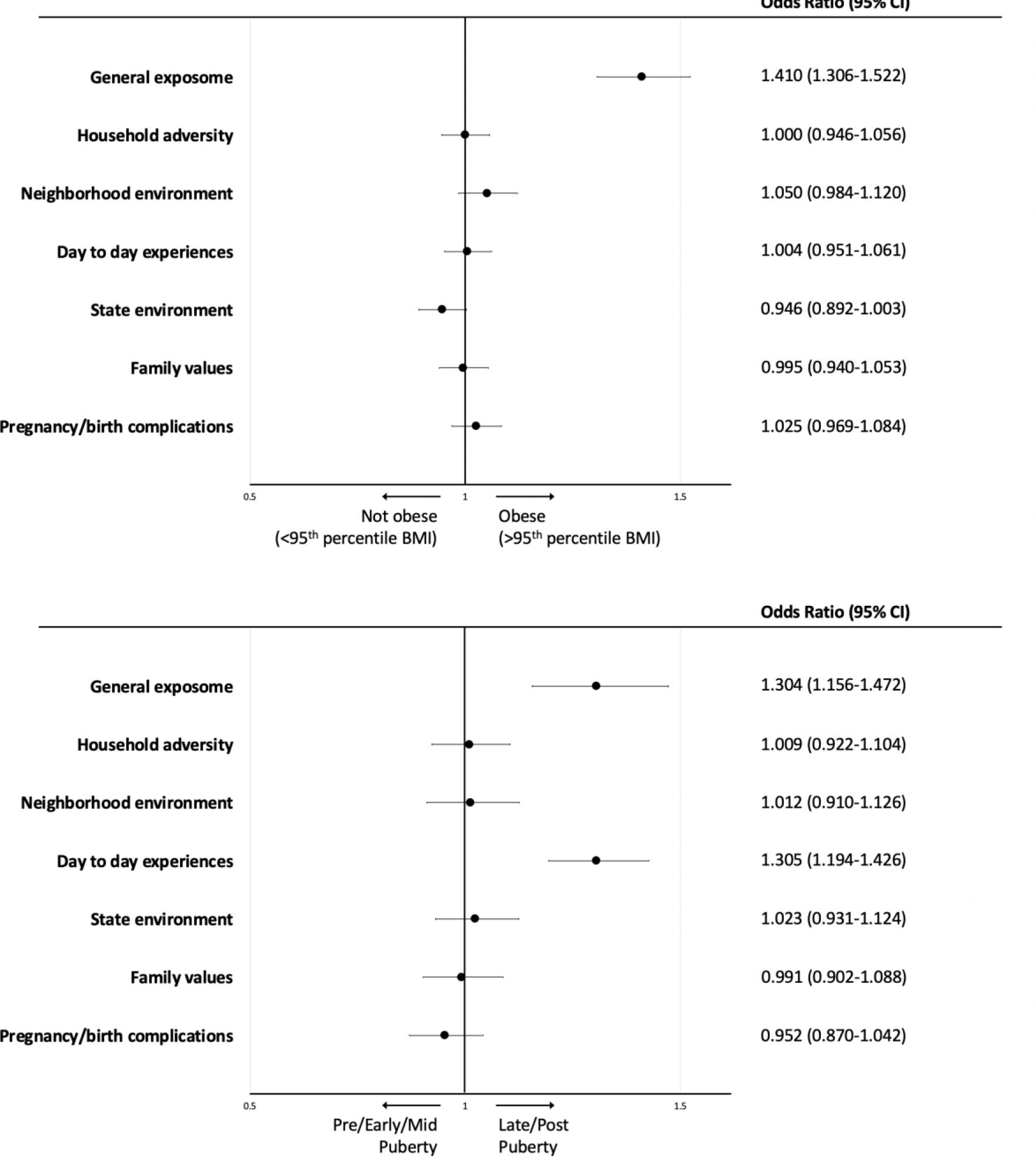
Association of exposome factor scores with obesity and pubertal development in the ABCD Study. Association of the exposome factor scores with obesity (binary variable, BMI>=95^th^ percentile, **top panel**) and late or post-pubertal stage (binary variable, contrasted against pre-, early, and mid-pubertal stage, **bottom panel**). Odds ratios were extracted from a binary logistic regression model with exposome scores as independent variables, covarying for age, sex, race (White, Black, Other), ethnicity (Hispanic), parent education, and household income. Puberty model also co-varies for BMI. Abbreviations: BMI = body mass index; 95% CI = 95% confidence interval.

### Generalization of the exposome framework in the Philadelphia Neurodevelopmental Cohort (PNC)

#### Generation of exposome scores in PNC

To test the generalizability of the exposome framework outside of the ABCD Study, we employed a confirmatory analytic approach in an independent US youth dataset – the PNC, which was sampled between 2009-2011^28^, more than 5 years before the onset of the ABCD Study. Notably, PNC was ascertained through a hospital network (Children’s Hospital of Philadelphia) and not school networks as in the ABCD Study. We age-matched the PNC generalization sample through limiting the age of PNC participants to under 14 years (the entire sample ranged from 8-21 years), resulting in a total of N=4,993 participants with a mean age of 10.9 years, like the ABCD Study sample. Except for age and similar gender distribution, the PNC sample displayed notable differences compared to the ABCD Study sample, including a greater proportion of Black participants (31.6% in PNC vs. 20.2% in ABCD) and a smaller proportion of Hispanic youths (7.3% in PNC vs. 20.1% in ABCD). Notably, PNC was a single site study (compared to 21 sites in ABCD). **Supplemental Table 15** details demographic characteristics of the generalization cohort and the corresponding measures in the ABCD Study.

For the PNC exposome analysis, we identified all available environmental exposures in the PNC (n=29 variables) and performed a bifactor confirmatory factor analysis of the exposures, with the goal of obtaining acceptable model fit. Indeed, fit of the model was acceptable^52, 53^, with a RSMEA of 0.036±0.001, SRMR of 0.068, and CFI of 0.94. This confirmed one portion of the exploratory ABCD analysis (also a bifactor model), but it also allowed us to generate orthogonal scores from the PNC model, including a “general exposome” score (as done in the ABCD sample). Notably, the generation of a PNC general exposome score allows one to test associations with mental and general health measures in the attempt to replicate findings from ABCD Study, despite that PNC had much “leaner” characterization of environment compared to ABCD Study (n=29 variables in PNC compared to n=798 variables in ABCD Study, with no data on school and family dynamics in PNC). As seen in **Supplemental Figure 2,** exposome factor analysis of all 29 environmental factors included four factors. *Factor 1* comprises variables that are broadly related to *household adversity* and include first degree family history of mental health issues and parental separation/divorce. *Factor 2* comprises variables most related to *neighborhood environment*, informed by census-derived measures. *Factor 3* comprises variables related to *trauma exposure*. *Factor 4* comprises two variables most related to *early life*, including birth complications and lead exposure. **Supplemental Table 16** details the environmental exposures in PNC and the loading on the exposome factors obtained from the confirmatory factor analysis.

#### Association of exposome scores with mental health, obesity and pubertal development in PNC

We then tested the capacity of PNC exposome factors in explaining variance in health using similar measures as in ABCD. For mental health, we used the *P-factor* (as in ABCD) that we had previously calculated based on the clinical assessment of PNC^63, 64^. For general health measures, we used obesity and being at a more advanced pubertal developmental stage (late/post puberty), as in the ABCD analyses.

Consistent with the ABCD analyses, we found that the addition of the exposome factors substantially increased the variance explained (adjusted R^2^) in the P-factor, from <4% (when relying on demographics alone) to 18.4%, with the *Exp-factor* similarly associated with the P-factor, though at a smaller effect size than in ABCD (Standardized Beta=0.15, 95%CI 0.26-0.3, P<0.001 in PNC vs. Standardized Beta=0.285 in ABCD, see **Table 3** for full model statistics). In the general health measures, similar to the main analyses in ABCD, the *Exp-factor* was significantly associated with obesity (OR=1.43, 95%CI=1.27-1.61, P<0.001 in PNC vs. OR=1.41 in ABCD); and with advanced pubertal development (estimated by scoring 4/5 on a 5 Likert scale of pubertal development, OR=1.26, 95%CI=1-1.59, P=0.047 in PNC vs. OR=1.3 in ABCD, **Figure 5** and **Supplemental Tables 17-18**).

**Figure 5.**
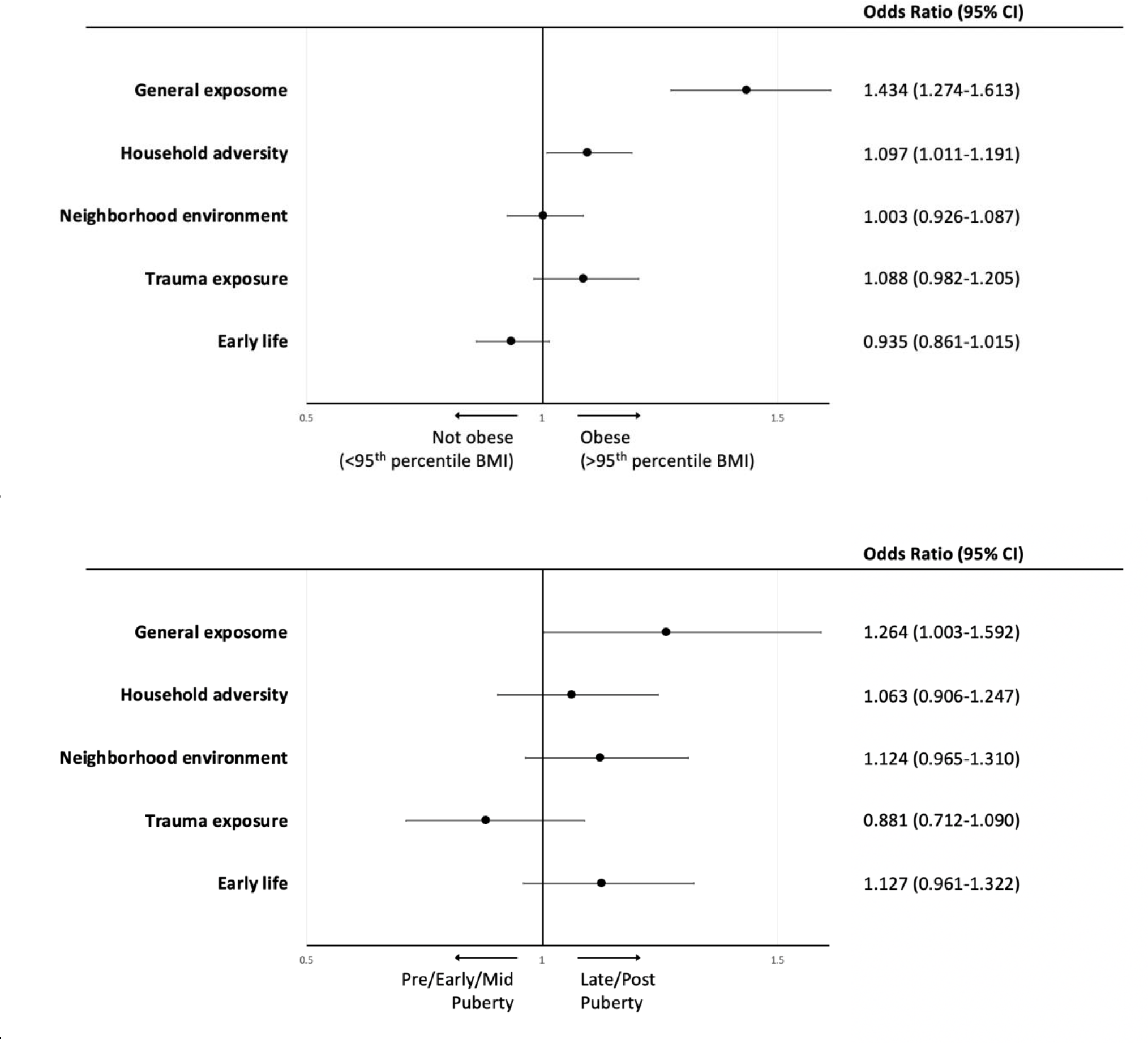
Association of exposome factor scores with obesity and pubertal development in the PNC. Association of the PNC exposome factor scores with obesity (binary variable, BMI>=95^th^ percentile, **top panel**) and late or post-pubertal stage (binary variable, contrasted against pre-, early, and mid-pubertal stage, **bottom panel**). Odds ratios were extracted from a binary logistic regression model with exposome scores as independent variables, covarying for age, sex, race (White, Black, Other), ethnicity (Hispanic), and parent education. Puberty model also co-varies for BMI. For models testing associations with pubertal measures, the PNC sample was limited to age range 10-12 to minimize age effects on models. Sample included N=1,496, of whom 271 were at late/post pubertal status. Abbreviations: BMI = body mass index; 95% CI = 95% confidence interval.

**Table 3.** Association of exposome factor scores to psychopathology *P-factor* score in the PNC.

|  | Model 1 (Demographics) |  |  | Model 2 (Demographics + Exposome) |  |  |
| --- | --- | --- | --- | --- | --- | --- |
|  | **Beta** | **SE** | **P** | **Beta** | **SE** | **P** |
| **Age (months)** | 0.029 | 0.02 | 0.05 | -0.024 | 0.02 | 0.085 |
| **Female sex** | -0.038 | 0.019 | 0.01 | -0.025 | 0.013 | 0.064 |
| **White race** | -0.120 | 0.026 | <0.001 | -0.067 | 0.024 | 0.006 |
| **Black race** | 0.009 | 0.028 | 0.736 | -0.012 | 0.025 | 0.640 |
| **Hispanic ethnicity** | -0.021 | 0.017 | 0.217 | -0.009 | 0.016 | 0.575 |
| **Parent education (years)** | -0.108 | 0.015 | <0.001 | -0.046 | 0.015 | 0.003 |
| **General exposome adversity** |  |  |  | 0.150 | 0.020 | <0.001 |
| **Household adversity** |  |  |  | 0.139 | 0.014 | <0.001 |
| **Neighborhood environment** |  |  |  | 0.025 | 0.014 | 0.072 |
| **Trauma exposure** |  |  |  | 0.314 | 0.018 | <0.001 |
| **Early life** |  |  |  | 0.091 | 0.014 | <0.001 |
| **Adjusted R<sup>2</sup>** | 0.039 |  |  | 0.184 |  |  |

Effect sizes (standardized betas) derived from a linear regression model testing association of demographics and exposome factors with general psychopathology (P-factor). Abbreviations: PNC= Philadelphia Neurodevelopmental Cohort. SE = standard error.

#### Sensitivity Analyses

We conducted multiple sensitivity analyses in ABCD to assess robustness of the main findings. We first aimed to test whether the association of exposome factors with mental health depends on the measure used to model mental health. In the main analyses, we modeled mental health dimensionally using that the P-factor, which is a reliable measure of psychopathology in youth samples^65, 66^ that represents life course vulnerability to psychiatric disorders^67^ and is predictive of long term psychiatric and functional outcomes^68^. In sensitivity analyses we tested associations of exposome factors with parent-reported child psychopathology available in ABCD (using the total child behavior checklist [CBCL] t-score). We found that similar to main analyses, addition of the exposome factors increased the explained variance by ∼7 fold to 17.8%, compared to 2.5% in the model relying on demographics, household income, and parent education (**Supplemental Table 19**). In addition, we also tested association of exposome factor with more clinically interpretable binary diagnoses based on the ABCD clinical assessment: depression and attention deficit hyperactivity disorder (ADHD). We chose these two diagnoses as they represent disorders of both internalizing and externalizing symptomatology. Results showed that, similar to models using dimensional psychopathology, exposome factors are associated with both depression and ADHD (**Supplemental Table 20**).

In sensitivity analyses of the general health measures, we tested association of exposome factors with continuous measures of weight (BMI percentiles) and puberty (1-5 Likert scale), rather than binary measures as in main analyses. Results were similar in direction and statistical significance to the main analyses **(Supplemental Tables 21-22)**.

In addition, we also conducted sensitivity analyses that accounted for potential site and family relatedness effects in ABCD. Because we wanted to evaluate environment based on factors that are included in the comprehensive exposome variable list (and not based on site), we did not account for ABCD Study sites in our main analyses. In sensitivity analyses we ran Mixed models testing the associations between exposome factors and mental health (P-factor and CBCL) and general health measures (BMI and pubertal development scale) accounting for site and family clustering. Results revealed similar findings as in main analyses **(Supplemental Tables 23-26)**, except for the anticipated loss of statistical significance of the *state-level environment* exposome factor effects (that depends on site since the ABCD Study included 21 sites from different states across the US). Notably, in the main analyses, clustering within site was intentionally not modeled because it was confounded with state-level variables. For example, if a state contained only one site, there would be no variability within that site on important state-level variables (e.g., cannabis legality). It is our working assumption that many of the quantitative ways that sites differ are accounted for by the state-level variables (indeed, that is their purpose).

Lastly, to maximize “harmonization” across ABCD and PNC datasets, we tested associations of exposome factors with P-factor, BMI, depression, ADHD, obesity and advanced pubertal status including identical co-variates that were available in both ABCD and PNC (age, sex, race, ethnicity, parental education). These analyses showed consistency across both youth cohorts (**Table 4**). Of note, in both studies, data were already collected, and analyses could not be truly harmonized, rather we tried to use similar measure as much as possible.

**Table 4.** Association of the general exposome factor score with health measures in ABCD Study and PNC using identical co-variates in both cohorts (age, sex, race, ethnicity, parental education).

| Cohort | Dependent variable | Standardized Beta | 95%CI | P |
| --- | --- | --- | --- | --- |
| ABCD | P-factor | 0.28 | 0.26-3 | <0.001 |
| PNC | P-factor | 0.15 | 0.11-0.19 | <0.001 |
| ABCD | BMI | 0.16 | 0.14-0.18 | <0.001 |
| PNC | BMI | 0.15 | 0.1-0.19 | <0.001 |
| Cohort | Dependent variable | Odds Ratio | 95%CI | P |
| ABCD | Depression | 1.61 | 1.44-1.80 | <0.001 |
| PNC | Depression | 1.28 | 1.06-1.46 | 0.012 |
| ABCD | ADHD | 1.35 | 1.27-1.43 | <0.001 |
| PNC | ADHD | 1.17 | 1.05-1.3 | 0.005 |
| ABCD | Obesity | 1.43 | 1.34-1.53 | <0.001 |
| PNC | Obesity | 1.43 | 1.27-1.61 | <0.001 |
| ABCD | Late/post pubertal | 1.37 | 1.23-1.52 | <0.001 |
| PNC<sup>a</sup> | Late/post pubertal | 1.26 | 1-1.59 | 0.047 |

Effect sizes derived from a linear regression model (standardized betas for continuous and odds ratio for binary measures) testing associations of the general exposome factor score with various health and mental health variables. Abbreviations: ABCD= Adolescent Brain Cognitive Development Study; PNC= Philadelphia Neurodevelopmental Cohort; BMI= Body mass index; ADHD= Attention deficit hyperactivity disorder.
<sup>a</sup>For the models in PNC testing associations with pubertal development status we included participants ages 10-12 (n=1,496).

## Discussion

We provide a comprehensive investigation of the exposome in early adolescence in the US in two separate large youth samples. We show that a data-driven approach allows integration of multiple exposures, resulting in dimensional factors representing different facets of the exposome, and that these factors explain variance in measures of early adolescent general and mental health. Our findings in ABCD Study, which was done in 21 sites across the US, allow for the appreciation of quantitative differences among American children’s environments across sociodemographic groups, which are associated with their trajectories of mental and physical development throughout the lifespan^69, 70^. Notably, a major finding is that, within orthogonal exposome subfactors, significant items loaded from different measurement tools and levels of analysis (parent- and youth-report, individual-level exposures, and census-derived variables).

This suggests that specific exposures within exposome factors likely represent a shared latent factor, highlighting the need to use a theoretical exposome framework when studying the relationship between environment and health^16^. Furthermore, bifactor modeling of the exposome revealed a general exposome adversity factor that was obtained independently in two separate cohorts, even though one cohort provided substantially more detailed environmental data than the other (n=798 exposures in ABCD and n=29 in PNC). This general exposome adversity factor integrates multiple exposures in addition to orthogonal exposome subfactors. While the current study analyzed cross-sectional data and cannot be used to infer causality, we suggest that our work provides a roadmap for dissection of environmental effects on developmental outcomes while accounting for the exposome’s complexity.

This research is important for several reasons. *First*, it demonstrates how inevitably collinear environmental exposures can be modeled when they are captured at multiple levels. For example, the *household adversity* subfactor in ABCD had strong loadings on youth-report of parental trouble with the law, parental self-reported psychopathology, developmental history (capturing prenatal exposure to cannabis), and parent-report of poverty and whether pregnancy was planned. Therefore, when trying to dissect associations of specific exposures with developmental outcomes based on *a priori* knowledge and hypotheses in the ABCD Study, one should account for the collinearity that is likely to confound any relationship that a specific exposure may have with an index outcome of choice. *Second*, our results suggest that data-driven approaches to characterize the exposome may be important to reveal latent factors that cannot be identified with *a priori* knowledge. A key example is the prenatal exposure items in ABCD, from which items split between the *household adversity* subfactor (prenatal exposure to substances, planned pregnancy) and the *pregnancy/birth complications* subfactor. Notably, growing efforts try to link pre-/post-natal exposures in the ABCD Study with developmental outcomes (prenatal cannabis exposure^33^, breastfeeding^71^ and other prenatal adversities^72^). Hence, it will become increasingly important to rigorously account for exposome complexity to allow generalizability and replicability of findings and identify causal mechanisms that are not confounded by collinear exposures. *Third*, in the context of understanding variance in psychopathology, our findings provide compelling evidence for the critical need to include environmental exposures when modeling psychopathology outcomes. We observed ∼ 5- to 10- fold increase in R^2^ explaining dimensional psychopathology upon addition of exposome factors when using different psychopathology measures in two independent cohorts, over and above the commonly used estimators of socioeconomic environment (parent education and household income). Of note, while we could not test for causality in this work, we suggest that the inclusion of exposome scores in predictive models of psychopathology (where causality is not the focus), may improve their performance considerably. *Fourth*, our finding on exposome contribution to variance in obesity and pubertal development in two independent samples provides a proof-of-concept for the utility of studying exposome effects on health trajectories in youth as they mature. *Fifth*, our ability to generalize the exposome framework and show that a general exposome factor can be calculated in an independent youth sample that is different in both its demographic characteristic and in its much leaner environmental phenotyping, may suggest that our findings have implications for modeling environmental effects in other developmental cohorts in the US and globally.

Previous research in other youth cohorts supports the notion that different exposures (e.g., trauma and neighborhood SES) and different mechanisms of environmental stress (threat vs. deprivation) are differentially associated with brain and behavior outcomes^73^, highlighting the need to address environmental complexity. For example, growing literature supports the notion that different exposures are associated with distinct brain structures and networks^37, 74^. The deep phenotyping of multiple environmental facets in the ABCD Study creates unprecedented opportunities to specifically link environmental effects to brain and behavior development.

Recent ABCD studies have provided proof-of-concept for brain-behavior-environment analyses that map neural parameters to multiple exposures^75, 76^, and for the potential to use a subset of environmental risk factors to explain variance in mental health outcomes^77^. In addition, several studies have reported associations of specific exposures with cognition and neuroimaging parameters in ABCD data (e.g., household income^42^, neighborhood disadvantage^78^, lead exposure^41^). The studies mentioned above all used baseline ABCD data, which does not include key environmental exposures. The current study expands on previous works as we used 1-year follow-up data, which included youth-reported exposures (negative adverse life events and experiences of discrimination) not captured at baseline. Notably, these items had high loadings on the *Exp-factor* and represent a total of 5 exposures among the top-loading 13. That the *Exp- factor* explains substantial variance in both mental and general health indicators emphasizes the need to incorporate youth-report when studying the exposome.

We suggest that this study be considered a roadmap when modeling environment in future investigations of developmental trajectories in longitudinal cohort studies. Notably, the current study does not investigate the exposome’s associations with cognitive and imaging measures, which could be done in future works utilizing multimodal and longitudinal datasets. Additionally, exposome scores can be used to explore interactions within the exposome (ExE), which have been identified in association with baseline ABCD cognitive and imaging outcomes^41^. Similarly, exposome scores can be used as covariates to adjust for nuisance environmental variance in studies with smaller samples or when trying to dissect the link between a specific exposure and an outcome. Moreover, we suggest that integration of genetic data with the exposome scores can facilitate better modeling when studying GxE mechanisms in developmental cohorts, allowing researchers to reliably measure environment (with all its complexities) as dimensional construct in conjunction with polygenic risk scores as dimensional genetic burden^79, 80^, as recently shown in an adult cohort^24^. Lastly, our findings in ABCD Study reveal large quantitative differences in latent environmental factors that illuminate disparities among demographic groups in America, which likely relate to disparities in later lifespan health outcomes^81^. We suggest that the exposome scores can be used to identify and focus on high-risk subgroups in large population cohorts that are more difficult to identify using *a priori* knowledge. Studies of such subpopulations are critical in the effort to tease apart mechanisms of resilience^82^, which are themselves influenced by multiple dimensions of environment (i.e., intrapersonal, family, neighborhood)^83^, and therefore require investigation in a wide environmental context.

A few methodological considerations we took are worth discussion. First, when determining environmental variables to include in analysis, we generally tried to take an inclusive approach informed by literature on environmental effects on development^2, 84, 85^. We included some variables that have substantial genetic components (e.g., parent psychopathology) and others that are confounded by psychopathology (e.g., school enjoyment). We chose not to include substance use variables, which we considered to reflect “psychopathology indicators” rather than environmental exposures in the young age range of this study. Second, we chose to use a bifactor model to fit the exposome data. This was largely in anticipation of a general exposome factor that would “absorb” any correlations among the latent factors. This model also produces orthogonal scores useful in downstream analyses to interpret specific effects. These decisions are rationalized and detailed in full in **Methods**.

## Limitations

Our findings should be viewed considering several limitations. First, we acknowledge that although we attempted to include all possible environmental factors in the two datasets, we nevertheless had to follow a reasoned decision-making process to determine what exactly to include in our analyses. For example, in ABCD we used composite scores as opposed to raw scores in some instances; and in PNC we chose to include specific geocoded Census variables based on our previous works. These decisions could have influenced results. Nevertheless, the current analysis provides, to our knowledge, the most comprehensive evaluation of environment in developmental cohorts and includes youth-report of key adversities that were not included in previous studies. Second, we used cross-sectional data to test associations of the exposome factors with psychopathology, obesity, and pubertal development. Longitudinal studies are warranted to evaluate temporal relationship between exposome and health trajectories and identify causal mechanisms. Third, there are inherent limitations to collection of environmental data, such as the retrospective report of events and recall bias. Fourth, our study does not address the complexity of genetic contribution to environmental exposures (including gene-environment correlations). This line of research is critical to address specificity of exposome effects on development and merits thorough future investigation outside the scope of the current work.

Fifth, while PNC was similar to the ABCD Study sample in terms of mean age and gender distribution, it was significantly different in its racial/ethnic composition and it had significantly fewer environmental exposures that we could use for replication. Relatedly, each dataset had its inherent limitations. PNC was done in one site, making it impossible to address state-level environment exposome. In contrast, a sample as complex as the ABCD Study, with its 21 sites, includes much potential for measurement invariance violations- for example, by race, by sex, by site, and other demographic groupings. It is important for future research to investigate consistency of measurement models across groups and sites, but it is beyond a scope of the current work. Finally, we did not take a “best practice” approach to the factor analyses (i.e., split the sample, estimate an EFA model in one portion, and test the EFA model in a CFA in the other portion). However, we did not intend to test a theoretical structural model, not even the one “found” by the EFA. Instead, the purpose was to derive scores from the model that most reasonably fit the entire ABCD and PNC datasets. We suggest that cross-validation of the scores will occur as they are used in downstream analyses, especially of longitudinal data that is and will be available for both cohorts.

## Conclusion

We leveraged two large, diverse datasets of US adolescents with deep phenotyping of environmental exposures to produce a roadmap for studying the exposome in youth. We propose that the exposome paradigm allows research to move beyond “looking under the lamp post” to a rounded dimensional investigation of environmental burden during development. We hope that future studies will build on the exposome framework in longitudinal cohorts to better understand developmental trajectories of youths through its integration in multi-omic research of brain, behavior, and health.

## Methods

### Participants

The ABCD sample includes 11,878 children aged 9–10 years at baseline, recruited through school systems^86^. For the purposes of this study, 1-year follow-up data was used (N=11,235).

Participants were enrolled at 21 sites, with the catchment area encompassing over 20% of the entire US population in this age group. All participants gave assent. Parents/caregivers signed informed consent. The ABCD protocol was approved by the University of California, San Diego Institutional Review Board (IRB), and was exempted from a full review by the University of Pennsylvania IRB. See **Supplemental Table 1** for full demographic data.

The PNC is a collaboration between the Children’s Hospital of Philadelphia (CHOP) and the Brain Behavior Laboratory at the University of Pennsylvania. Participants were from the greater Philadelphia area were ascertained through the Children’s Hospital of Philadelphia (CHOP) pediatric health care network. The PNC included children aged 8–21 years (N = 9, 498). For participants aged 8–10, clinical evaluation including probing for suicidal ideation was done using a parent report. For participants 11 and older, clinical evaluation was based on an interview with the youth. For the current study, to keep with the developmental stage of the ABCD sample, we only included PNC participants under age 14 years old (N=4,933, see **Supplemental Table 15** for demographic data). Participants’ written assent and parental consent were obtained.

University of Pennsylvania and CHOP’s Institutional Review Boards approved all procedures.

### ABCS Study analyses

#### Measures

We included a total of 798 variables that tap participants’ environmental exposures at multiple levels of analysis including family-, household-, school-, extracurricular-, neighborhood-, and state-level, as well as prenatal exposures. We included measures based on both youth- and parent-report, as well as geocoded address. We did not include genetic data as we focused on environmental exposures in this project. Additionally, we did not include imaging or neurocognitive data. Imaging procedures and the comprehensive ABCD Study neurocognitive assessment were not conducted in the ABCD Study time point used in the current exposome analysis (i.e., the 1-year follow-up assessment). **Supplemental Table 2** provides the full range of exposure measures used in the present study.

For the models testing associations of exposome scores with psychopathology (*P-factor*), we used variables tapping mental health (n=93, see **Supplemental Table 27** for the full list) comprising youth self- or caregiver-reported attitudes, experiences, and problems. For models testing the exposome’s association with obesity and pubertal development, we used BMI and pubertal development data (measure pds_y_ss_female_category_2 and pds_y_ss_male_cat_2).

All measures were collected at the ABCD 1-year follow-up assessment.

### Statistical Analysis

The analytic plan and hypotheses were preregistered on Open Science Framework in October 2020, before the full release of ABCD 1-year follow-up data. Analyses were conducted from January to October 2021, following ABCD data release 3.0, which was the first full release of the 1-year follow-up data and included youth-reported life events and discrimination. We used R^87^ (package psych^88^) and Mplus 8.4^89^ for factor analyses and SPSS statistical package version for all other statistical methods. Statistical significance was set at P<0.05.

### Handling of missing data

Models testing associations of the exposome with psychopathology, obesity, and pubertal development used listwise deletion of missing data. All other analyses use pairwise deletion.

### Dimensionality reduction of environment in ABCD analyses

Due to the large number of ABCD variables of multiple formats (continuous, ordinal, and nominal) and from multiple measures (youth-report scales, parent-report scales, census-level composites, etc.) of different lengths (scales used in the ABCD Study ranged from 2 to 59 items in length), the process of arriving at an optimal ABCD exposome model was complex.

**Supplemental Figure 1** presents a visual schematic of the steps taken to reduce dimensionality of variables. We started with 798 variables, from which we selected certain ABCD-provided summary variables according to a combination of *a priori* knowledge (e.g. similar decisions had to be made about the American Community Survey in our previous works^38^) and common sense, ultimately collapsing variable count to 348. We often chose to use summary scales to represent overarching culture and environment (e.g., Mexican American Cultural Values Scale, family conflict) and indicators of health (e.g., family psychiatric history, dietary habits). We included these in the following analysis and, using multiple exploratory factor analyses (EFAs), iteratively reduced the number of variables. We elaborate below, but the first iteration is representative of later iterations. It proceeded as follows:

1. Estimate a mixed correlation matrix where each bivariate relationship in the matrix is appropriate to the variable types. If two variables are continuous, use a Pearson correlation; if they are both dichotomous, use a tetrachoric correlation; if they are both ordinal (or one ordinal and one dichotomous), use polychoric; if one is continuous and the other dichotomous, use biserial; and if one is continuous and the other ordinal, use polyserial.
2. Determine the number of factors to extract based on subjective evaluation of the plot of descending eigenvalues (scree plot). That is, visually, subjectively determine where on the scree plot the decreasing function begins to form a linear trend (find the “elbow”). **Supplemental Figure 3** shows an example of a scree plot for determining the number of factors to extract.
3. Estimate an EFA model using least-squares extraction and oblimin rotation.
4. Examine the solution for interpretability, with particular attention to groups of variables so strongly related that they should be reduced. For example, if a factor comprised items from only one scale, with very high loadings on that factor and near-zero loadings elsewhere, that would suggest the scale could be reduced.
5. Use secondary factor analyses to reduce the groups of variables discovered in #4 above. For example, if all items from a checklist of negative life events loaded together in the solution in #4 above, submit that checklist to its own factor analysis. As in the main analysis, choose the number of factors based on subjective evaluation of the scree plot, calculate the appropriate correlation matrix (if a yes/no checklist, tetrachorics would be used), and use least-squares extraction with oblimin rotation.
6. Reduce the variables from #4 and #5 above by creating composite scores. In the present study, these composites were calculated using the following rules: a) if variables are dichotomous, take the mean to get a proportion endorsed; b) if variables are ordinal, z-transform them and take the mean; c) if variables are continuous, calculate factor scores (oblique Thurstone/regression method) from the model in #5 above.
7. Replace the variables discovered in #4 above with the variables created in #6 above. Using this updated data set, go back to #1 and repeat.

In the present study, the above steps were repeated 9 times (**Supplemental Tables 3-11**) to arrive at a set of 96 variables with minimal redundancy. Next, we estimated an EFA solution using the “clean” 96-variable dataset obtained from the iterative process described above. A unique aspect of this step was that, because we expected complex structure whereby some cross-loadings would be substantial and meaningful, we used iterated target rotation (ITR)^90, 91^ rather than a simple structure rotation like oblimin or promax. Whereas simple structure rotations attempt to get p-1 elements in each row as close to zero as possible (where p = number of factors), ITR allows salient cross-loadings to be estimated freely. It starts with a simple structure rotation (here, oblimin), uses the resulting pattern matrix to determine not only which item loads where but also which cross-loadings might be non-negligible, and builds a partially-specified target matrix that incorporates cross-loading items^92^. Specifically, it uses a user-defined threshold (here, 0.20), sets all elements of the target matrix at 0 for items loading below that threshold, and sets all other (non-negligible) loadings to “unspecified” (indicating they should be estimated freely). The results of this target rotation are then used in the same way as the original simple structure rotation to specify a new target, and the process is repeated. When a new target matrix matches a previous target matrix in the iterative process, the ITR solution has converged.

With the EFA solution obtained from the above ITR process, we went on to define a quasi- confirmatory bifactor analysis from which ABCD exposome factor scores could be obtained.

The bifactor model confirmatory factor analysis (CFA) was estimated in Mplus using the wlsmv estimator, accounting for clustering by family. A bifactor model uses a factor configuration whereby each variable loads not only on its specific factor (e.g., a measure of family poverty might load on a “household adversity” factor), but also on a general exposome factor comprising (with estimated loadings on) all variables. Note that this analysis reduced the included items from 96 to 65 according to significance of within-factor association. A visual presentation of the exposome bifactor solution is presented in **Figure 2**. Additionally, please see **Supplemental Table 28** for bifactor indices^93^, such as explained common variance (ECV), omega-hierarchical, and factor determinacy. Some aspects of our approach are unique and require clarification.

First, it is important to state why we used a CFA on the same sample as was used for the EFAs, whereas it’s typical to perform EFAs on a training sample to provide a configuration that CFA can then confirm in a separate sample. If we wished to make a claim about the “true” theoretical structure of the exposome, then a cross-validation framework would be optimal, as we have done in a previous work^94^. However, we conceptualize the exposome here as a bottom-up collection of phenomena that define it (the exposome) *ad hoc*. If additional variables were added to the analysis (e.g., prevalence of venomous snakes in the area or affordability of local fresh vegetables), the definition of the exposome itself would change. This is in contrast to, for example, depression, whose definition does not change when indicators are added; additional indicators simply increase the precision of measurement. In this sense, the goal of the present study was simply to calculate scores for use in downstream analysis (as shown in this study with the exposome factors’ association with psychopathology, obesity, and pubertal development), and confirmatory bifactor modeling allowed optimal estimation of those scores. Furthermore, it is important to clarify why a confirmatory model was used to calculate scores as opposed to the original, exploratory model. CFA was used here because, as of this study, there is no good bifactor rotation available. The most common “bifactor” rotation, the Schmid-Leiman, is not a true bifactor. It estimates a higher-order solution and transforms that to a bifactor configuration, which necessitates proportionality constraints on the solution. Another option is the Jennrich- Bentler true bifactor rotation ^47^, which has been shown to perform poorly in multiple studies to date^46^. It is therefore preferred to use a confirmatory bifactor model to obtain scores.

A second aspect of our approach that requires explanation is the decision to use a bifactor model at all, given the weak inter-factor correlations found in the final EFA (see **Results**). Bifactor modeling accounts for inter-factor correlations by modeling the overall factor as its own phenomenon, unlike, for example, orthogonal EFA rotations (like varimax), which force orthogonality onto solutions without accounting for the true obliqueness of the phenomena. Usually, one of the indications that a bifactor model might be useful is moderate-to-strong inter- factors correlations, which suggest the existence of an overall, general factor underlying all item responses^51^. Here, inter-factor correlations were weak, suggesting that there may not be a hierarchical structure to environmental exposures (neither second-order nor bifactor). However, in addition to common sense suggesting that adverse environments at the distal level beget adverse environments at the proximal level, there is increasing evidence that bifactor general factors can contain critically important information even when inter-factor correlations are weak^95^. This is possible because, while the subfactors of a model might correlate only weakly, individual items within each subfactor may still load strongly on the general factor. The above-cited example demonstrates not only that such a phenomenon exists, but that the general factor scores generated from the seemingly ill-advised models have substantial validity.

### Association of exposome scores with demographic characteristics

For comparisons of exposome scores within each demographic variable (males vs. females, high vs. low parent education and household income, and comparisons across race and ethnicity), we used t-tests (Bonferroni corrected for seven comparisons), with Cohen’s d to estimate effect size.

### Generation of P-factor in ABCD

The exposome analyses required some special modeling due to the mixture of variable formats (continuous, ordinal, etc.) and expected complex structure. By contrast, because all psychopathology variables (n=93) in this study were items (youth self- or caregiver-reported attitudes, experiences, and problems; see **Supplemental Table 27** for the full variable list), they could be analyzed entirely within an item-factor analysis framework^96^ whereby all correlations are polychoric rather than being a mix of types. This analysis (using oblimin rotation) revealed that the psychopathology items clustered exactly by instrument (i.e., questionnaire/scale), with only two cross-loadings >0.30; see **Supplemental Table 29**). The “clean” solution supports our use of a simple structure rotation. All items thusly grouped by instrument form a 6-factor solution. Specifically, *Factor 1* comprises variables most related to symptoms of psychosis and associated prodrome. *Factor 2* comprises variables most related to suicidal ideation or attempt (suicidality). *Factor 3* comprises variables most related to externalizing symptoms. *Factor 4* comprises variables most related to manic symptoms. *Factor 5* comprises variables most related to self-reported (mostly internalizing) symptoms. *Factor 6* comprises variables most related to positive affect.

The results of the configuration above were taken as the basis of the confirmatory model used to calculate the *P-factor* score using a bifactor model CFA estimated in Mplus using the wlsmv estimator, accounting for clustering by family. **Supplemental Table 30** details results from confirmatory bifactor model analysis, displaying specific factor loadings as well as loadings to a general psychopathology factor. Overall, fit of the model was acceptable (CFI=0.93; RMSEA=0.023; SRMR=0.085), and these results are presented visually in **Supplemental Figure 4**. This general *P-factor*score was used for subsequent correlational analyses with the exposome factor scores.

### Associations of exposome scores with the mental health in ABCD

We tested the association of exposome scores (the general *Exp-factor* and the six orthogonal subfactors) with the *P-factor* (dependent variable in the main analysis) and with total CBCL t-score (in sensitivity analysis) using a linear regression with the seven exposome factors as independent variables and age, sex, parent education, household income, race (White, Black, Asian, Other), and Hispanic ethnicity as covariates. The model was also run without the exposome scores to estimate the change of adjusted R^2^ upon addition of exposome scores to the model.

### Association of exposome scores with obesity and pubertal development in ABCD

We tested the association of exposome scores (the general *Exp-factor* and the six orthogonal subfactors) with obesity or pubertal development (two separate models) using a binary logistic regression model with obesity (binary variable, BMI percentile>=95); or with advanced pubertal development status (binary variable of being late/post-pubertal stage [4/5 on a 5 Likert scale of pubertal development] contrasted against pre-/early-/mid-pubertal status [1-3 on the Likert scale]) as the dependent variables, and the seven exposome factors as independent variables, co-varying for age, sex, parental education, household income, race (White, Black, Other), and Hispanic ethnicity. The pubertal development model also co-varied for BMI.

### Sensitivity analyses

We conducted sensitivity analyses where we used other mental health measures as dependent variables instead of the P-factor, as in main analyses. We ran linear regression models with the total child behavior checklist [CBCL] t-score and binary logistic regression models with binary diagnosis of depression or ADHD based on the K-SADS interview. In all of these models, exposome factors were the independent variables with the same co-variates as in main analyses (age, sex, race, ethnicity, household income and parent education).

We also conducted sensitivity analyses for the models of general health measures, using binary logistic regression with continuous BMI percentile scores and continuous pubertal scale as dependent variable, instead of binary measures (obesity and advanced pubertal status) as in main analyses. In both models, exposome factors were the independent variables and co-variates were identical to main analyses (age, sex, race, ethnicity, household income, parent education, and BMI as an additional covariate in the model of pubertal development).

Lastly, to account for clustering within site and family, we ran mixed-effects regression models for both mental health (with P-factor and CBCL scores) and general health measures (with BMI and pubertal scales) with random intercepts for site and family using the lmer() function in the lmerTest package.

### PNC analyses

#### Measures

Lifetime history of psychopathology symptoms were evaluated by trained and supervised Bachelor’s and Master’s level assessors who underwent rigorous standardized training and certification using a structured screening interview ^28^, based on the Kiddie Schedule for Affective Disorders and Schizophrenia (K-SADS) ^97^. Generation of P-factor scores was done as described above for the ABCD Study, using item-wise (i.e., symptom-level) psychopathology responses (total 110 items) from the clinical interview across all assessed psychopathology domains^63, 64^.

#### Generation of Exposome scores in PNC

To generate exposome scores, we assembled all environmental variables that were collected as part of the PNC assessment. As in the ABCD Study, we used a permissive definition of environment and considered family history of psychiatric disorders as an environmental exposure. The exposures included (i) family history of psychiatric disorders based on the abbreviated version of the Family Interview for Genetic Studies (Maxwell, 1996) and an indicator whether the parents are separated or not; (ii) traumatic experiences assessed with a screener for eight traumatic experiences (yes/no items) that fulfill criterion A in post-traumatic-stress-disorder diagnosis^36^; (iii) census neighborhood (block-group-level) measures derived from participants’ geocoded address ^38^; (iv) two items tapping early life exposure: birth complication and history of lead exposure (both binary yes/no items).

Generation of the PNC exposome score was done using a confirmatory bifactor model, generating a general (adversity) PNC exposome score and four subfactors. Fit of the model was judged based on the same indices as described above for the ABCD portion (CFI, RMSEA, and SRMR). Additionally, please see **Supplemental Table 28** for bifactor indices^93^, such as explained common variance (ECV), omega-hierarchical, and factor determinacy.

#### Association of exposome scores with psychopathology, obesity and puberty in PNC

After the generation of the exposome scores, we followed the same approach as in the ABCD Study and tested the association of exposome scores with the P-factor (linear regression) with obesity (BMI percentile>95%) or advanced pubertal development (binary logistic regression). In the models testing associations with pubertal development, we limited the PNC sample to ages 10-12 (n=1,496), to minimize the large age effect sizes that were present when using age range 8-13 on pubertal development in the full PNC generalization sample. Models co- varied for age, sex, race (White, Black, Other), Hispanic ethnicity and parental education.

#### “Harmonized” models across ABCD and PNC

In attempt to maximize similarity across the two datasets, we ran similar regression models (linear for continuous measures and binary logistic for binary measures) with exposome factors as independent variables co-varying for measures that were available in both ABCD and PNC: age, sex, Race, Hispanic ethnicity, and parental education (**Table 4**).

## Data Availability

Data used in the preparation of this article were obtained from the Adolescent Brain Cognitive Development Study (https://abcdstudy.org), held in the National Institute of Mental Health Data Archive. A full list of supporters is available at https://abcdstudy.org/federal-partners.html. A listing of participating sites and a complete listing of the study investigators can be found at https://abcdstudy.org/consortium_members/. Data preprocessing and analysis are detailed at https://github.com/barzilab1/abcd_exposome.

https://nda.nih.gov/abcd

https://www.ncbi.nlm.nih.gov/projects/gap/cgi-bin/study.cgi?study_id=phs000607.v3.p2

## Acknowledgements

Data used in the preparation of this article were obtained from the Adolescent Brain Cognitive Development^SM^ (ABCD) Study (https://abcdstudy.org), held in the NIMH Data Archive (NDA). This is a multisite, longitudinal study designed to recruit more than 10,000 children age 9-10 and follow them over 10 years into early adulthood. The ABCD Study® is supported by the National Institutes of Health and additional federal partners under award numbers U01DA041048, U01DA050989, U01DA051016, U01DA041022, U01DA051018, U01DA051037, U01DA050987, U01DA041174, U01DA041106, U01DA041117, U01DA041028, U01DA041134, U01DA050988, U01DA051039, U01DA041156, U01DA041025, U01DA041120, U01DA051038, U01DA041148, U01DA041093, U01DA041089, U24DA041123, U24DA041147. A full list of supporters is available at https://abcdstudy.org/federal-partners.html. A listing of participating sites and a complete listing of the study investigators can be found at https://abcdstudy.org/consortium_members/. The Philadelphia Neurodevelopmental Cohort was supported by grants RC2MH089983, RC2MH089924.

## Author Contribution

TMM and RB conceptualized and designed the study, conducted the analyses, interpreted the data and drafted the first version of the manuscript. EV, STA, GED, IS, JW and AN substantially contributed to study design, organization and analysis of data and visualization of findings, and have all substantially contributed to revision of the first draft of the manuscript. RCG, REG, VW, and SG substantially contributed to study design and conceptualization, interpretation of findings, and have provided substantial input to revision the manuscript from its first draft till its final version. All authors have approved the submitted version and have agreed to be accountable to the submitted study.

## Disclaimer

ABCD consortium investigators designed and implemented the study and/or provided data but did not participate in analysis or writing of this report. This manuscript reflects the views of the authors and may not reflect the opinions or views of the National Institutes of Health or ABCD consortium investigators.

